# First and Second Wave Dynamics of Emergency Department Utilization during the COVID-19 Pandemic: a retrospective study

**DOI:** 10.1101/2021.12.09.21267520

**Authors:** Robi Dijk, Patricia Plaum, Stan Tummers, Frits H.M. van Osch, Dennis G. Barten, Gideon H.P. Latten

## Abstract

**Background:** Since the COVID-19 pandemic, there has been a decrease in emergency department(ED) utilization. Although this has been thoroughly characterized for the first wave(FW), studies during the second wave(SW) are limited. We examined the changes in ED utilization between the FW and SW, compared to 2019 reference periods.

**Methods:** We performed a retrospective analysis of ED utilization in 3 Dutch hospitals in 2020. The FW and SW (March-June and September–December, respectively) were compared to the reference periods in 2019. ED visits were labeled as (non-)COVID-suspected.

**Findings:** During the FW and SW ED visits decreased by 20.3% and 15.3%, respectively, when compared to reference periods in 2019. During both waves high urgency visits significantly increased with 3.1% and 2.1%, and admission rates (ARs) increased with 5.0% and 10.4%. Trauma related visits decreased by 5.2% and 3.4%. During the SW we observed less COVID-related visits compared to the FW (4,407 vs 3,102 patients). COVID-related visits were significantly more often in higher need of urgent care and ARs where at least 24.0% higher compared to non-COVID visits.

**Interpretation:** During both COVID-19 waves ED visits were significantly reduced, with the most distinct decline during the FW. ED patients were more often triaged as high urgent and the ARs were increased compared to the reference period in 2019, reflecting a high burden on ED resources. These findings indicate the need to gain more insight into motives of patients to delay or avoid emergency care during pandemics and prepare EDs for future pandemics.

**Funding:** None

## Background

From November 2019 onwards, coronavirus disease (COVID-19) spread rapidly around the world and in March 2020, the World Health Organization (WHO) declared the outbreak a pandemic.^1^ In response to the steep increase of COVID-19 cases hospital organizations prepared for increasing patient volumes by changing workforce and infrastructure with particular focus on emergency departments (EDs).^2^

In the Netherlands, the first COVID-19 diagnosis was confirmed on February 27^th^, 2020. Government regulations were increased stepwise until a partial lockdown was imposed on March 23^rd^. This included the closure of public institutions (such as schools) and stay at home policies for those with possible COVID-19 symptoms.

At the end of the first wave (FW, March to June 2020) in total 50,273 citizens had been infected by COVID-19, 11,397 patients had been admitted to a hospital (2,939 (25.8%) to an intensive care unit (ICU)), and 10,067 people had died due to COVID-19 in the Netherlands.^3–5^

In July and August 2020, most measures were lifted after a rapid drop in COVID-19 infections and hospital admissions. By September, however, infection rates started to spike again. As a consequence, a second lockdown was enforced.^6^ During the second wave (SW, September to December 2020) in total 726,314 people have been infected. In the SW, 19,354 patients were admitted to a hospital (3,629 (18.8%) to an ICU), and 10,046 people died from a COVID-19 infection.^3,4^ In the early stages of the pandemic, citizens were advised to only seek care when absolutely necessary.^7^ Concerns about non-COVID-19 emergencies were subsequently raised when a 30% reduction in ED visits was observed.^8,9^ It was hypothesized that this reduction was due to both COVID-19 fears and lockdown effects, including reduced exposure to injury-prone activities, improved hygiene measures in the community and downscaling of non-acute healthcare.^10^ Other studies found that a relatively large proportion of ED patients reported delay in seeking emergency care.^11,12^ During the SW, elective-non-COVID-care was continued as long as hospital capacity allowed for. In contrast to the FW, studies on ED utilization during the SW are limited.

As the pandemic continues and infection rates keep arising, it is of utter importance to gain insight in population needs and subsequent emergency care. Only then we can deliver appropriate care and future health care related damage can be minimized.

In this retrospective observational study, we investigated ED utilization and patient volumes during the FW and SW of the COVID-19 pandemic in the Netherlands, examined differences between those waves and compared with pre-COVID reference periods.

## Methods

### Design and setting

In this retrospective observational study, we investigated the utilization of 3 hospital-based EDs in the southeast of the Netherlands during the first and second wave of the COVID-19 pandemic (from March-June and September–December 2020, respectively). Identical periods in 2019 were used as a reference. The 3 EDs combined serve a population of 760,000 individuals. Annual census is 35,000 patients for ED 1, 25,000 for ED 2 and 25,000 for ED 3.

### Patients

All patients who visited one of the EDs during the study period were eligible for inclusion.

Patients were excluded when it was unclear whether the ED visit was COVID-19 related and when patients refused participation in retrospective studies. In addition, we excluded patients in which ED length of stay was likely to be documented incorrectly, either 0 minutes or longer than 10 hours were considered to be outliers.

### Analysis and statistics

We retrieved information from standard digital patient records: age, gender, date of ED visit, possible COVID-19 infection, triage urgency (using the Manchester Triage System, MTS)^13^, ED length of stay, referral route, admission and whether the ED visit was trauma related or not. Patients triaged red or orange using the MTS are classified as high urgency visits (seen by doctor within 10 minutes). Yellow, green and blue triage colours (seen by doctor within 1-3 hours) are classified as low urgency visits. From April-May 2020, the EDs prospectively labelled patients as COVID-19 suspected using the criteria in table 1. We retrospectively retrieved the missing labels for possible COVID-19 infection in patients who presented before hospitals had implemented prospective tracking, using identical criteria (Table 1).

**Table 1:**
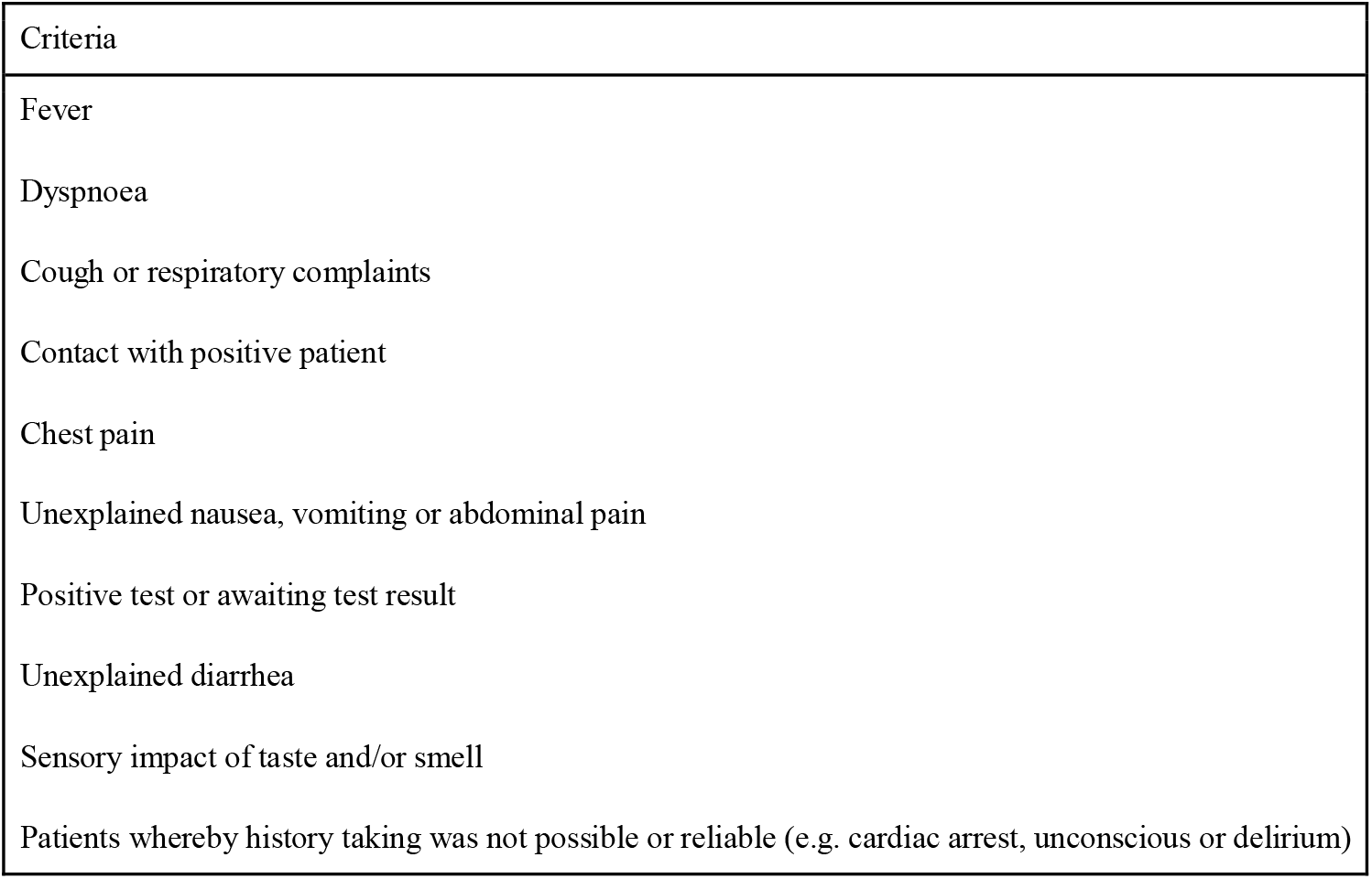
Criteria for possible COVID-19 infection

| Criteria |
| --- |
| Fever |
| Dyspnoea |
| Cough or respiratory complaints |
| Contact with positive patient |
| Chest pain |
| Unexplained nausea, vomiting or abdominal pain |
| Positive test or awaiting test result |
| Unexplained diarrhea |
| Sensory impact of taste and/or smell |
| Patients whereby history taking was not possible or reliable (e.g. cardiac arrest, unconscious or delirium) |

**Table 1:** overview of overall ED utilization during COVID waves and reference periods in 2019.

|  | First wave | Reference period first wave | Second wave | Reference period second wave | p value first wave vs reference | p value second wave vs reference | p value first vs second wave |
| --- | --- | --- | --- | --- | --- | --- | --- |
| Total number of patients, n | 18,204 | 22,607 | 18,282 | 21,557 | <0.001 | <0.001 | 0.608 |
| Age in years, median (IQR) | 62 (39-76) | 59 (32-75) | 62 (37-76) | 60 (33-76) | <0.001 | <0.001 | <0.011 |
| Sex |  |  |  |  |  |  |  |
| Female, n (%) | 8,722 (48.4%) | 11,242 (49.7%) | 8,867 (48.5%) | 10,697 (49.7%) | <0.001 | <0.001 | 0.754 |
| Male, n (%) | 9,298 (51.6%) | 11,363 (50.3%) | 9,415 (51.5%) | 10,856 (50.3%) | <0.001 | <0.001 | 0.866 |
| Possible COVID, n (%) | 4,777 (26.5%) | 0 (0%) | 3,102 (17.0%) | 0 (0%) | <0.001 | <0.001 | <0.001 |
| Transportation to ED: Ambulance |  |  |  |  |  |  |  |
| Self-referral, n (%) | 3,801 (21.1%) | 3,716 (16.7%) | 4,053 (22.2%) | 3,745 (17.8%) | 0.811 | <0.007 | <0.044 |
| GP, n (%) | 2,924 (16.3%) | 3,110 (14.0%) | 2,744 (15.0%) | 3,035 (14.4%) | 0.125 | <0.002 | 0.126 |
| Transportation to ED: Own transport |  |  |  |  |  |  |  |
| Self-referral, n (%) | 877 (4.9%) | 1,402 (6.3%) | 966 (5.3%) | 1,287 (6.1%) | <0.001 | <0.001 | 0.231 |
| GP, n (%) | 8,324 (46.2%) | 11,192 (50.4%) | 8,079 (44.2%) | 10,353 (49.1%) | <0.001 | <0.001 | 0.301 |
| Other, n (%) | 2,081 (11.6%) | 4,222 (19.0%) | 2,432 (13.3%) | 2,675 (12.7%) | <0.001 | <0.009 | <0.001 |
| Urgency of ED visit* |  |  |  |  |  |  |  |
| high urgency, n (%) | 3,823 (22.9%) | 4,247 (19.8%) | 3,817 (22.6%) | 4,169 (20.5%) | <0.001 | <0.001 | >0.999 |
| low urgency, n (%) | 12,847 (77.1%) | 17,125 (80.2%) | 13,043 (77.4%) | 16,182 (79.5%) | <0.001 | <0.001 | 0.686 |
| Trauma, n (%) | 4,933 (27.4%) | 7,375 (32.6%) | 5,251 (28.7%) | 6,916 (32.1%) | <0.001 | <0.001 | <0.004 |
| Median minutes spent in ED, m (IQR) | 157 (105-219) | 147 (94-208) | 154 (99-215) | 147 (96-207) | <0.001 | <0.001 | <0.001 |
| Admission after ED visit, n (%) | 8,990 (57.3%) | 10,490 (52.3%) | 8,674 (56.7%) | 10,146 (46.3%) | <0.001 | <0.001 | 0.218 |

If the total number of patients does not match the total of inclusions, this is due to missing value.
\* Using the MTS, red or orange triage colour are classified as High urgency (seen by doctor <10 minutes) and triage colour yellow, green or blue are classified as low urgency (1-3 hours before assessment).
IQR = interquartile range

We compared the ED utilization between the FW and SW and between 2020 and their respective reference periods in 2019.

Data were analyzed using IBM Corp. Released 2020. IBM SPSS Statistics for Macintosh, Version 27.0. Armonk, NY: IBM Corp. Descriptive analyses were used for patient characteristics. Continuous data were reported as means with standard deviation (SD) and compared using Students’ T test, or as medians with interquartile ranges (IQR) and compared using the Mann Whitney U test. Categorial data was reported as absolute numbers and as valid percentages (to correct for missing data); they were compared using chi-square or Fisher exact tests. A p-value <0.05 was considered statistically significant.

The study was approved by the medical ethical committee of Zuyderland Medical Center, Heerlen, the Netherlands (METCZ20210031).

## Results

56,719 patients visited one of the three EDs in 2020, compared to 65,938 patients in 2019. Of all included patients, the median age was 60.5 years (IQR 34-76) and 50.7% was male. In total, 9,034 (16.0%) of the ED patients in 2020 were labelled as possible COVID-19 infected, of which 4,777 presented during the FW and 3,102 during the SW (Figure 1). In total, 127,060 patients were eligible for inclusion, 4,403 (3.5%) were considered outliers and were excluded (4,362 0-minute visits and 41 more than 10 hours).

**Figure 1:**
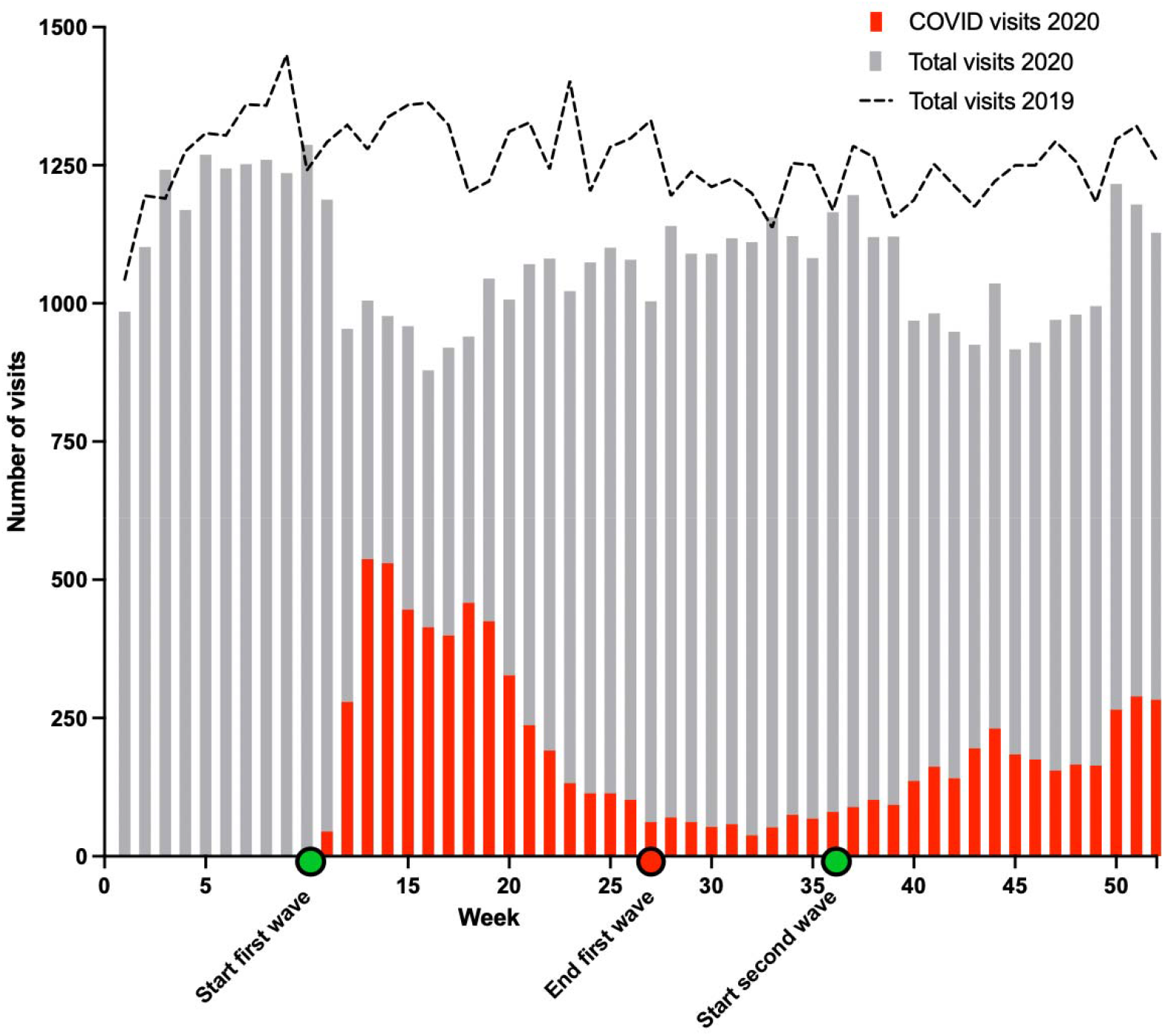
Overview of patients visiting the ED over 2019 and 2020.

### First wave compared to second wave

During the uprise of COVID-19 in the FW there was a decline of 20.3% in patients presenting to the ED, compared to a less distinct decline during the SW of 15.3% (both compared to 2019) as shown in Figure 1. When comparing the FW and SW, the number of patients with possible COVID-19 infections was higher during the FW (4,777, 26.5%) compared to the SW (3,102, 17.0%, p<0.001). Furthermore, an increase in trauma related visits (27.4% vs 28.7%, p<0.001) and a decrease in admission rates (AR) during the SW was observed (57.3% vs 56.7%, p<0.001).

### Comparison between 2020 and 2019

The dynamics of ED utilization during the two waves and their respective reference periods in 2019 are shown in Table 2. Patients transported to the ED by ambulance increased by 6.6% (p<0.001) and 5.0% (p<0.001) in the FW and SW, respectively. Furthermore, the percentage of patients triaged as high urgency increased by 3.1% in the FW (p<0.001) and 2.1% in the SW (p<0.001). Trauma related visits decreased by 5.2% in the FW (p<0.001) and 3.4% in the SW (p<0.004). Finally, AR increased respectively by 5.0% (p<0.001) and 10.4% (p<0.001) during the first and second pandemic wave.

**Table 2:**
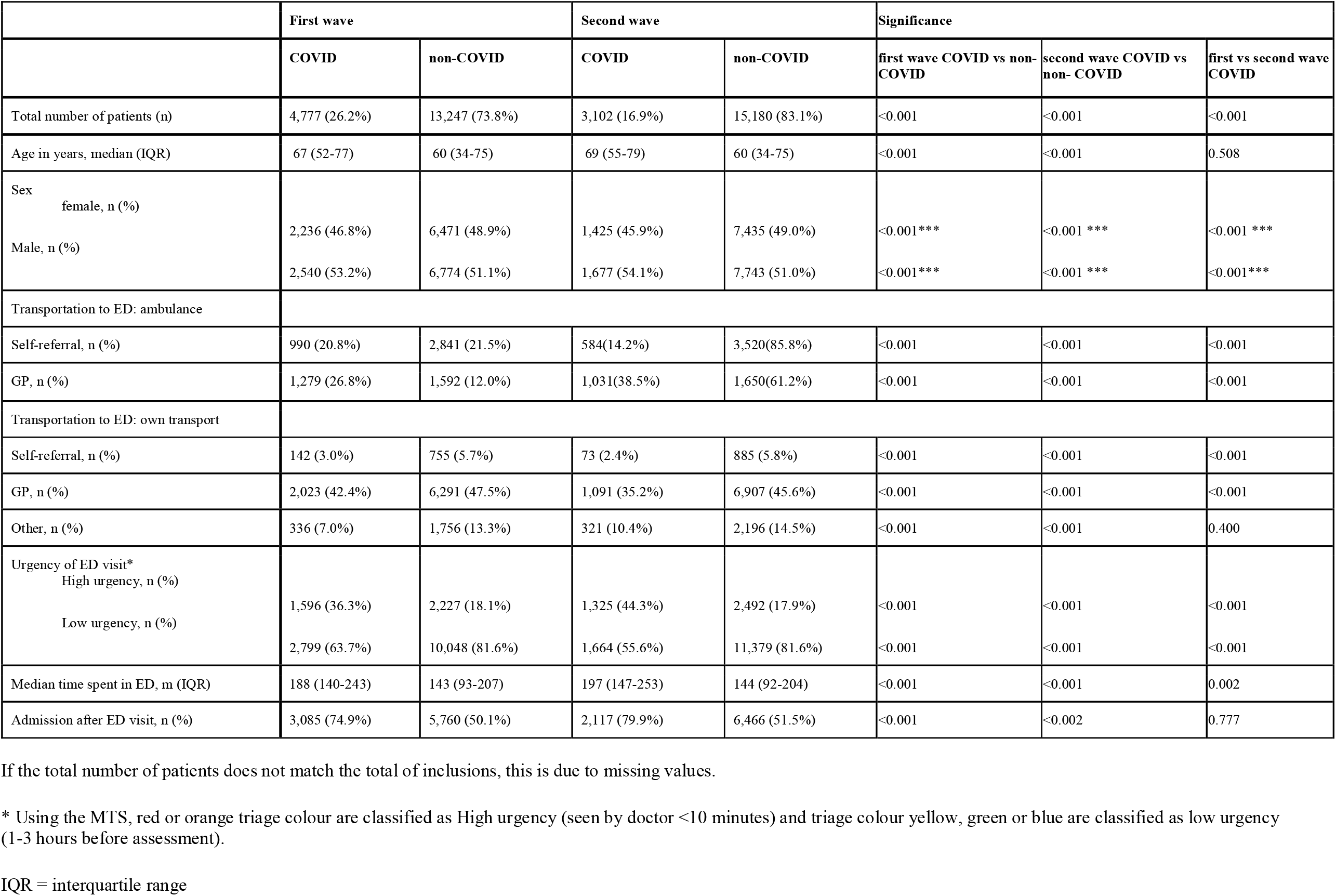
comparison of ED utilization between possible COVID vs non-COVID ED visits during first and second wave.

### Possible COVID-19 versus non-COVID-19

Table 3 depicts a comparison between possible COVID-19 vs non-COVID-19 visits during the FW and SW. High urgent visits were 18.2% (p<0.001) and 26.4% (p<0.001) more common in patients with possible COVID-19 compared to non-COVID-19 visits during the first and second wave, respectively.

During the first and second wave, the AR for suspected COVID-19 patients was significantly higher at 74.9% and 79.9% compared to 50.1% (p<0.001) and 51.5% (p<0.001) for non-COVID-19 suspected patients.

## Discussion

During the first two COVID-19 waves in the Netherlands, a significant decrease in the number of patients presenting to the ED was observed, with the most distinct decline during the FW. When comparing the two waves with their reference periods in 2019, there was an increase in transportation by ambulance, more patients were triaged as high urgent and there were higher ARs, particularly in COVID-19 suspected patients.

Furthermore, a decline in trauma-related ED visits was observed. The increased triage urgency and ARs may reflect a higher workload of EDs during the pandemic waves in year 2020.

Previous studies corroborate our results and also report a significant drop in ED visits, increase in high urgent visits and a decline in trauma-related visits during both waves.^7–9,11,13–15^ However, to our knowledge, this is one of the first studies comparing ED utilization during the first and second wave as well as (non-)COVID-19 related visits.

The reduced ED utilization during the two pandemic waves could be attributed to several possible causes, including pure lockdown effects, fear and uncertainties about a novel infectious disease, and misperceptions about the accessibility of EDs.^11^ The lockdowns, which included the closure of non-essential shops, offices and the leisure industry, were associated with lower mobility rates and less traffic or workplace accidents.^15^

The decline of ED utilizations was more prominent during the FW than during the SW. This may be explained by a reduction of ‘viral fear’, the increasing knowledge about the virus and repeated encouragements by the government as well as healthcare organizations to seek care when necessary.^7^ This observation might also be explained by the rebound effect of delayed healthcare seeking behavior, possibly leading to an exacerbation of the neglected pathological conditions and healthcare damage. Furthermore, the measurements taken by the government were less strict, which probably resulted in higher mobility rates and increased traffic, explaining the increase of trauma related visits.^12^ Finally, during the SW more elective, non-COVID care was continued. This may have been associated with higher rates of patients who suffered from complications, both postoperatively and medication related.

During the SW, ED utilization still was significantly lower compared to the reference period, possibly resulting from modified community spread of (viral) infections caused by the lockdown, social distancing and other measures during the pandemic.^14^ This conception is reinforced by a highly unusual respiratory syncytial-virus outbreak in July-August 2021 after loosening of the lockdown measures.^16^

The strengths of this study include its multicenter design, great number of patients included, mostly prospective labeling of (non-)COVID-suspection and preclusion of selection bias by including all patients presenting to the three EDs. Limitations include the retrospective nature of this study, prospective labeling may have been different between hospitals, the COVID waves continued past the end of our study period and 9,790 patients (17.3%) have been labeled retrospectively because there was no prospective labeling at the very beginning of the pandemic.

Similar FW and SW dynamics were observed in all three EDs, based in a country with a well-developed primary care system and relatively low numbers of self-referrals. As a result, this study adds to the body of literature about ED utilization during the global COVID-19 pandemic. Future studies should focus on characteristics of the third and possible following waves of this ongoing pandemic. Furthermore, it would be desirable to gain more insight into motives of patients to delay or avoid emergency care.

## Conclusion

During both COVID-19 waves ED visits were significantly reduced, with the most distinct decline during the FW. ED patients were more often triaged as high urgent and the ARs were increased compared to the reference period in 2019, reflecting a high burden on ED resources. These findings indicate the need to gain more insight into motives of patients to delay or avoid emergency care during pandemics and prepare EDs for future pandemics.

## Data Availability

All data produced in the present work are contained in the manuscript

## About the Authors

Department of Emergency Medicine, VieCuri Medical Center, Venlo, Netherlands (Drs. Barten); Department of Emergency Medicine, Zuyderland Hospital, Heerlen & Sittard-Geleen, Netherlands (Drs. Latten) and Department of Clinical Epidemiology, VieCuri Medical Center, Venlo, Netherlands (Dr. van Osch).

Correspondence and reprint requests to Frits H.M. van Osch, Department of Clinical Epidemiology, VieCuri Medical Center, PO Box 1926, 5900 BX Venlo, Netherlands.

## Conflict of interest Statement

The authors have no conflicts of interest to declare.

### Abbreviation

AR: admission rate
COVID-19: coronavirus disease
ED: emergency department
FW: first wave
GP: general practitioner
IQR: interquartile Range
MTS: Manchester Triage System
PPE: personal protective equipment
SARS-CoV-2: severe acute respiratory syndrome coronavirus 2
SW: Second wave
WHO: World Health Organization

## Data sharing statement

1. Will individual participant data be available (including data dictonaries)? No
2. What data in particular will be shared? Not available
3. What other documents will be available? Not available
4. When will data be available? Not applicable
5. With Whom? Not applicable
6. For what types of analyses? Not applicable
7. By what mechanism will data be made available? Not applicable

## Appendix

A graph showing Google mobility trends during the COVID waves. ^15^

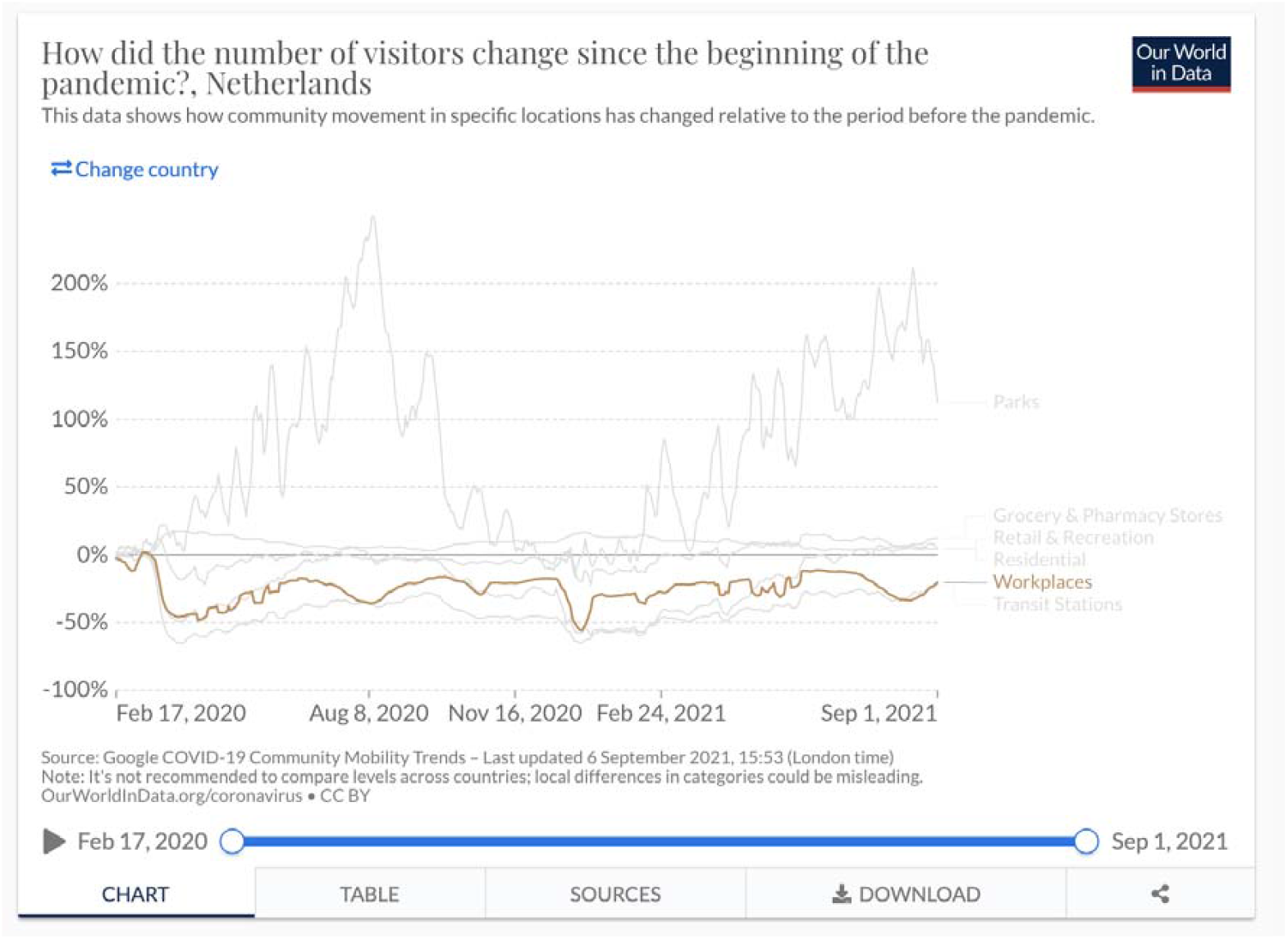

